# Plasma oxylipin levels may predict Covid-19 patient outcomes. An observational and retrospective study

**DOI:** 10.1101/2021.07.21.21260954

**Authors:** Denise Biagini, Maria Franzini, Paolo Oliveri, Tommaso Lomonaco, Silvia Ghimenti, Andrea Bonini, Federico Vivaldi, Lisa Macera, Laurence Balas, Thierry Durand, Camille Oger, Jean-Marie Galano, Fabrizio Maggi, Alessandro Celi, Aldo Paolicchi, Fabio Di Francesco

## Abstract

**Background:** The key role of inflammation in the progression of COVID-19 was evident right from the beginning of the pandemic. Consequently, many authors studied the onset and evolution of the cytokine storm. In contrast, other inflammatory mediators such as oxylipins have received almost no attention.

**Methods:** We conducted a monocentric observational and retrospective study (IMPRE-COVID-19) with patients (aged ≥ 18 years) admitted to Pisa University Hospital with COVID-19 related pneumonia. SARS-CoV-2 infection was confirmed by polymerase chain reaction in a nasopharyngeal swab, while pneumonia was demonstrated by CT scan. Oxylipin plasma levels were analysed in a convenient sample of 52 patients randomly selected from those that had a complete set of cytokine values evaluated for clinical purposes. In 12 cases, plasma samples collected on different days were available at the BMS Multispecialistic Biobank of Pisa University Hospital and were analysed to understand the evolution of the oxylipin levels during hospitalization. The two datasets that included oxylipin and cytokine values were analysed by principal component analysis (PCA), computation of Fisher’s canonical variable, and a multivariate receiver operating characteristic (ROC) curve.

**Findings:** Between March and April 2020, 52 patients were enrolled of which 28 were hospitalised in COVID-19 wards, 20 in ICUs, and 4 who were initially hospitalised in the wards and who were then transferred to the ICUs due to deteriorating health conditions. Plasma samples collected on different days were available from 7 patients in the wards, 1 from an ICU, and 4 from patients who were hospitalised in both. The mean age of participants was 61 years (SD 16), 11 were females (21%), and 41 were males (79%), without significant differences between groups in terms of age and gender. PCA of oxylipin data led to a clear differentiation of samples collected in COVID-19 wards and ICUs. This differentiation had not been obtained with cytokine data. In addition, borderline samples were from patients hospitalised in COVID-19 wards that were about to be transferred to the ICUs, thus suggesting that differentiation is not a consequence of a different treatment, but somehow related to a diverse evolution of the pathology. Computation of Fisher’s canonical variable identified the original input variables that were the most effective in discriminating between the two classes. The combination of the two datasets did not improve the discrimination, thus suggesting that oxylipins are more informative than cytokines for this purpose. A multivariate class model built using the four lowest-order principal components as the input variables, and COVID-19 ward samples as the target class, produced a ROC curve with a resulting area under the curve (AUC) equal to 0·92 – which is much higher than most AUC outcomes obtained for individual oxylipins.

**Interpretation:** After analysing the metabolic pathways of the most informative oxylipins, we speculate that more severe COVID-19 is associated with a selective deficiency of pro-resolving oxylipins leading to ineffective resolutive mechanisms of inflammation, likely worsened by endothelial damage. We believe that our oxylipin data suggest the possibility to predict the evolution of COVID-19 in individual patients at an early stage.

**Funding:** Institutional funds from the University of Pisa supported the study.

## Introduction

Approximately 20 months after the onset of the COVID-19 pandemic, despite the unprecedented effort of the scientific community, there are still many open questions regarding the pathophysiology of the disease, therapy, and the clinical management of patients. Around 15% of patients undergo severe COVID-19 and require hospitalization with oxygen supplementation, and 3–5% of these are admitted to intensive care units (ICUs) for ventilation support.^1^ A critical care triage is required to prioritize patients for intensive care and to optimize the use of limited resources,^2^ however unfortunately current physiological prediction scores are inaccurate.

We speculated that markers associated with acute immune-inflammatory response and severe COVID-19 symptoms could be found among chemicals involved in the cytokine and oxylipin storm.

We then realized that oxylipins could provide clinicians with clear indications regarding the urgency of patient transfer to ICU.

The cytokine storm results in a detrimental dysregulation of T cell responses and in an uncontrolled overproduction of immune cells and cytokines, such as tumour necrosis factor-α (TNF-α), interferon-γ (IFN-γ), interleukins (IL-1β, IL-2, IL-6, IL-7, IL-8), and a large number of chemokines (CCL2, CCL3, CCL5, CCL9, CCL10).^3^ Besides cytokines, other important inflammatory mediators, e. g. oxylipins, sustain the inflammatory response observed in severe COVID-19 cases.^4–8^ Oxylipins are bioactive lipids generated from both ω-3 and ω-6 polyunsaturated fatty acids (PUFAs) through enzymatic (e.g. prostanoids, epoxy and hydroxy fatty acids) and non-enzymatic (e.g. isoprostanoids) oxidation reactions.^9,10^

Because of their anti-inflammatory action, ω-3 PUFAs seem to limit the level and duration of the critical inflammatory phase^11^, and reduce ICU admissions of COVID-19 patients.^12^ At the same time, PUFAs act as precursors for pro-inflammatory, anti-inflammatory, and specialized pro-resolving lipid mediators (SPM).^13,14^ Firstly, pro-inflammatory mediators such as prostaglandins, thromboxanes, and leukotrienes are released into the body, leading to the classic signs of inflammation.^15^ The production of oxylipins then undergoes lipid mediator class switching mainly because of CYP450-derived epoxy fatty acids, thus shifting from the lipoxygenase pathway to the specialized pro-resolving mediators.^15^ SPMs mostly consist of lipoxins, resolvins, maresins, and protectins, which could lower the inflammatory response and even promote its resolution^16,17^ without being immunosuppressive unlike classic anti-inflammatory drugs.^18^

Consequently, a better understanding of the cytokine storm and its coupling with the oxylipin storm should provide new insights into the complex immuno-inflammatory cascade induced by SARS-CoV-2 and lead to novel strategies for managing patients.

In this paper, we compare the plasma levels of 48 oxylipins and 5 cytokines in COVID-19 ward and ICU patients and we show that the oxylipin plasma levels can be used to discriminate between the two classes by multivariate analysis.

## Research in context

### Evidence before this study

A key role of the cytokine storm in COVID-19 progression was suggested from many authors at an early stage of the pandemic. We hypothesized that oxylipins, another class of inflammatory mediators, could be involved. We searched PubMed and Google Scholar for articles published up to 15 July 2021, using the search items “lipid mediators in COVID-19”, “cytokine storm (or oxylipin storm) in COVID-19”, or “lipid mediators as predictors of COVID-19 severity”. Several review articles suggested that the role of both the cytokine and oxylipin storm should be investigated in relation to COVID-19 onset and progression, but we only found three papers and one preprint concerning explorative studies of lipid mediator levels in COVID-19. Among them, three manuscripts (Archambault *et al*., McReynolds *et al*., and Pérez *et al*.) revealed higher levels of lipid mediators in COVID-19 patients compared to healthy subjects, whereas Bosio *et al*. suggested a lipid dysregulation from moderate to severe disease. However, neither clear lipidome dysregulation nor statistically significant separation between the two groups was observed. We found no papers that highlighted an overall lipid mediator deficiency in severe versus moderate COVID-19 patients or that presented a panel of oxylipins for differentiating those patients that required ICU assistance from the others.

### Added value of this study

To our knowledge, this is the first study to suggest that oxylipins may be heavily involved in the inflammatory response to SARS-CoV-2 and represent possible biomarkers for differentiating COVID-19 patients based on severity. We found that ICU patients showed a lower (down to two orders of magnitude) plasma concentration of anti-inflammatory (EETs, HETEs) and pro-resolving mediators (D-resolvins, maresins, protectins), thus suggesting the importance of targeting the lipid mediator class switching for a timely picture of a patient’s ability to respond to the viral attack. Moreover, the fact that four patients that had been hospitalised in medical wards and appeared as borderline subjects in the PCA plot had to be transferred to the intensive care unit within 1-4 days after sample collection, suggests that a model exploiting selected lipid mediators as biomarkers might be able to identify patients at risk of severe COVID-19 and a negative outcome.

### Implications of all the available evidence

The striking difference between concentrations of several oxylipins observed in COVID-19 wards and ICU plasma samples may help to shed new light on the pathophysiological mechanism of COVID-19. We feel that these neglected chemicals deserve more attention. Our findings also suggest that new insights into the evolution of the pathology can be gained by studying the metabolic pathways that produce these inflammatory mediators and the associated biological processes. A prediction model that classifies patients based on their risk of developing severe COVID-19 and having a negative outcome would prompt clinicians to act in order to increase patients’ chances of survival and optimize the use of resources. Not all the implications are clear at this stage, but they may well become apparent in the future in a wider range of viral pneumonias.

## Methods

### Study design and participants

This monocentric observational and retrospective study (IMPRE-COVID-19) was approved by the local Ethics Committee. Patients eligible for enrolment were aged 18 years or older and admitted to the University Hospital of Pisa (Pisa, Italy) in March-April 2020 with COVID-19 related pneumonia. SARS-CoV-2 infection was confirmed by polymerase chain reaction in a nasopharyngeal swab, while pneumonia was demonstrated by CT scan. Members of the Pisa COVID group collected all the clinical information, including co-morbidities, drug intake, food supplements and routine laboratory data, which were de-identified and stored according to the recommendations of the Ethics Committee and made available to academic researchers. Residuals of plasma samples used for routine clinical measurements were de-identified and stored in the BMS Multispecialistic Biobank of Pisa University Hospital.

Oxylipin levels were analysed in a convenient sample of 52 patients randomly selected among those with a complete set of cytokine values evaluated for clinical purposes. In a few cases, plasma samples collected on different days were available at the biobank and were analysed to understand the evolution of the oxylipin levels during hospitalization.

### Procedures

#### Virological tests

Viral nucleic acids were manually extracted from 250 μL of plasma or serum samples using the EXTRA blood kit (ELITechGroup, Turin, Italy) according to the manufacturer’s instructions. After extraction, purified RNA samples were screened by RT-qPCR using the SARS-CoV-2 R-Gene assay (Biomerioux, Marcy-l’Etoile, France) on an ABI 7500 FAST thermocycler (Applied Biosystems). The real-time SARS-CoV-2 R-Gene assay is carried out by two triplex PCRs. The first PCR detects the N gene and the RdRp gene. The second PCR detects the E gene of the SARS-CoV-2 genome. The assay contains internal controls to check PCR processing, and a cellular control to check sampling for certain results.

### Analytical procedures

#### Sample collection and processing

Peripheral blood samples were collected from COVID-19 ward and ICU patients with COVID-19 during their hospitalization, as part of the routine clinical activity of the Laboratory of Clinical Pathology of Pisa University Hospital. Whole blood was collected in EDTA tubes (BD Vacutainer^®^) and centrifuged at 1500 g for 10 minutes at 25°C to separate blood cells and plasma. Plasma was removed and stored in aliquots at -80°C. For the oxylipin analyses, the antioxidant butylhydroxytoluene (BHT, 15 mg/mL in methanol) was added before storage (1:100, BHT:sample) to preserve polyunsaturated fatty acids from in vitro lipid peroxidation. Before the analysis, a preliminary check identified specimens containing active SARS-CoV-2 virus, which were excluded from the quantification of the oxylipin content for safety reasons.

#### Quantification of oxylipins

The MS-based targeted profiling of 60 oxylipins (e. g. prostaglandins, lipoxins, protectins, resolvins, hydroxy- and epoxy-fatty acids, F_2_-isoprostanes, F_3_-isoprostanes, F_2_-dihomo-isoprostanes, and F_4_-neuroprostanes) was performed using micro-extraction by packed sorbent (MEPS) liquid chromatography tandem mass spectrometry (MEPS-LC-MS/MS) platform.^19,20^ Before MEPS extraction, plasma proteins were precipitated by the sequential addition of salts (i. e. 250 µL of CuSO_4_· 5 H_2_O 10% w/v and 250 µL of Na_2_WO_4_ · 2 H_2_O 12% w/v) and acetonitrile (500 µL) to the plasma sample (500 µL). The supernatant was then diluted (1:6 v/v) with water. Compounds were quantified by calibration curves plotting the analyte to an internal standard peak area ratio versus the corresponding concentration ratio. Table S1 lists the targeted oxylipins and the main analytical parameters. Table S2 reports the oxylipin concentration levels in COVID-19 ward and ICU samples.

#### Quantification of cytokines

Plasma cytokines (i. e. IL-6, IL-1β, IL-10, TNF-α, CCL2) and granulocyte-macrophage colony-stimulating factor (GM-CSF) were quantified by automated ELISA assays according to the manufacturer’s instructions (see supplementary information). Table S3 reports the concentration levels in COVID-19 ward and ICU samples.

### Outcomes

The two coprimary outcomes were the observation of extremely different plasma oxylipin concentration levels (up to two orders of magnitude) in samples collected from patients hospitalised in COVID-19 wards and ICUs, and that a multivariate unequal class model (UNEQ) differentiated items of the two groups with a high level of efficiency (AUC 0·92 in the test set). The secondary outcomes were the identification of the most effective oxylipins in discriminating between the two classes, and the formulation of hypotheses connecting the observations and biological processes related to the evolution of the inflammatory response to COVID-19.

### Statistical analysis

Datasets D1 and D2, which include the plasma levels of 48 oxylipins (Table S2) and 5 cytokines (IL-6, IL-1β, IL-10, TNF-α, and CCL2, Table S3), respectively, were obtained from the analyses of 70 samples collected from patients hospitalised in COVID-19 wards (patients = 32, samples = 43) and ICUs (patients = 24, samples = 27) (four patients were in both groups). Twelve out of sixty oxylipins (Table S1) were excluded from D1 as the concentrations were below the limit of quantification for more than 50% of samples. A decimal logarithmic transform was used to correct for asymmetry characterising all the variables.^21^ Samples were randomly split into a training (n = 56) and a test set (n = 14): the former was used to build the models and the latter to independently estimate performances and consistency.

Data were analysed by a multivariate exploratory method (principal component analysis, PCA).^22^ Principal components (PCs) are orthogonal linear combinations of the original input variables aligned in the directions of maximum variability in the same orthogonal space. Under the hypothesis that variability entails information, PCs are ordered based on decreasing associated variance and importance, so that projection of samples onto the plane of the two lowest-order PCs enables a plot to be drawn that shows the most informative undistorted visualization of multivariate data and enabling the identification of patterns. As coordinates of samples on the PCs are called scores, this plot is called a score plot, while coefficients representing contributions of the original input variables to the PCs are called loadings.^23^

In the present study, PCA was applied after column autoscaling – a pre-processing transform that ensures the same importance a priori is given to all variables, irrespectively of their magnitude.^24^ Fisher’s canonical variable was then computed in the plane described by the two lowest-order PCs, as the direction that maximises the ratio between inter-class and intra-class variances.^25^ This axis represents the most discriminant direction, and its loadings, multiplied by the loadings of the PCs considered, indicate the importance of the original input variables in the differentiation of the classes. Finally, a multivariate receiver operating characteristic (ROC) curve was obtained by varying the confidence level of unequal class models and computing, at each step, sensitivity and specificity.^26– 28^ Multivariate data processing was performed by in-house Matlab routines (The MathWorks, Inc., Natick, USA, Version 2019b).

### Role of the funding source

The funders of the study had no role in the study design, data collection, data analysis, data interpretation, or writing of the report. All the authors had full access to all data and shared the final responsibility for the decision to submit for publication.

## Results

Between 1 March and 30 April 2020, 52 patients were admitted to Pisa University Hospital, 28 in COVID-19 wards, 20 in ICUs and 4, initially hospitalised in COVID-19 wards, who were then transferred to the ICUs due to deteriorating health conditions. The clinical characteristics, comorbidities and outcome of patients are reported in Table 1.

**Table 1.** Demographic and clinical baseline characteristics of enrolled patients from COVID-19 wards (W) and intensive care units (ICU). Data are represented as median (first and third quartile). Statistics: Student’s t-test (difference between means) on log-transformed data.

|  | W (n=32) | ICU (n=20) | p |
| --- | --- | --- | --- |
| **Comorbidities** |  |  |  |
| Diabetes (n; %) | 1; 3 | 5; 25 |  |
| Hypertension (n; %) | 10; 31 | 7; 35 |  |
| COPD/asthma (n; %) | 3; 9 | 0; 0 |  |
| Hypercholesterolemia (n; %) | 4; 13 | 4; 20 |  |
| Heart disease (n; %) | 13; 41 | 3; 15 |  |
| **Clinical characteristics** |  |  |  |
| Age, years (mean±SD) | 60 (51 – 78) | 63 (57 – 73) | 0.7700 |
| P/F admission, mmHg | 313 (263 – 377) | 256 (207-327) | **0.0215** |
| P/F nadir, mmHg | 182 (95 – 288) | 96 (72 – 118) | **0.0055** |
| SOFA score | 2.0 (1.0 – 3.0) | 2.0 (1.8 – 3.3) | 0.3724 |
| Creatinine, mg/dL | 0.98 (0.80 – 1.27) | 1.13 (0.83 – 1.46) | 0.4274 |
| While blood cells, cells·10<sup>3</sup>/μL | 5.9 (4.5 – 7.8) | 8.0 (6.9-11.9) | **0.0117** |
| Neutrophils, cells·10<sup>3</sup>/μL | 3.8 (2.9 – 5.8) | 6.1 (3.7 – 8.3) | 0.0623 |
| Lymphocytes, cells·10<sup>3</sup>/μL | 1.1 (0.7 – 1.3) | 0.6 (0.5 – 1.1) | **0.0309** |
| C-reactive protein, mg/dL | 5.7 (2.8 – 11.0) | 8.8 (3.1 – 13.6) | 0.4470 |
| **Outcome** |  |  |  |
| Survived (n; %) | 27; 84 | 17; 85 |  |
| ETI (n; %) | 0; 0 | 12; 60 |  |

Baseline characteristics were generally similar between the two groups, with the exception of the Horowitz index for lung function (P/F ratio) that identified the acute hypoxemic respiratory condition of ICU patients, as well as their leucocytosis and lymphocytopenia, which were related to the higher inflammatory response and more severe viral infection.

The plasma concentrations of oxylipins and cytokines are reported in Table S2 and Table S3, respectively. A separation of samples collected from COVID-19 wards (blue symbols) and ICUs (red symbols) is clearly visible in the oxylipin score plot (Figure 1a), but which is not observed in the case of cytokines (Figure 1b). This pattern is consistent for items of both the training set (full symbols), used to build the model and calculate PCs, and the test set (empty symbols), which were simply projected onto the PC plane^21^ for validation purposes.

**Figure 1.**
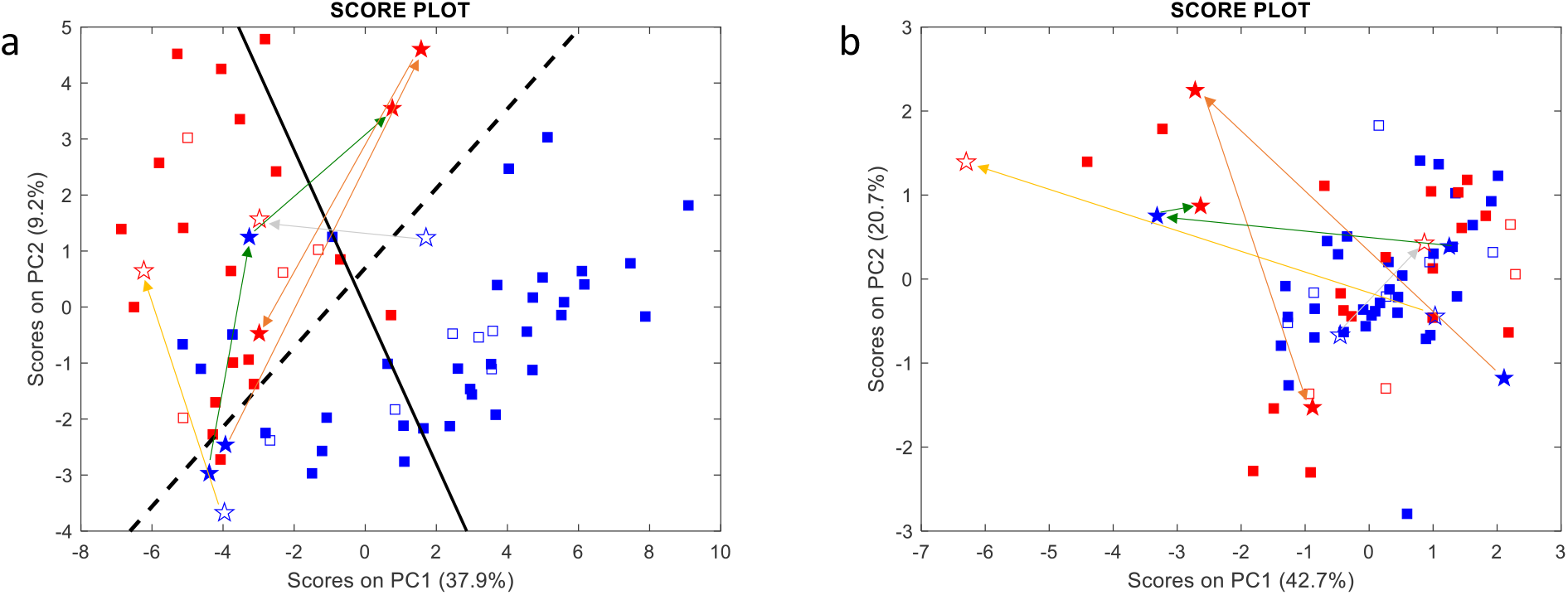
Oxylipin (a) and cytokine (b) score plots. Blue and red symbols represent samples collected from COVID-19 wards and ICUs, respectively, whereas full and empty symbols belong to the training and test sets, respectively. Samples collected from the same patient on different days are represented as stars connected by coloured arrows. The full black and dashed lines represent Fisher’s canonical variable, i.e., the direction of maximum discrimination between the two classes, and the delimiter separating the two classes, respectively.

Interestingly, many borderline samples were collected from COVID-19 ward patients who were transferred to the ICUs a few days after sample collection due to their deteriorating health conditions. Samples collected from the same patient on different days (stars) are connected by coloured arrows: shifts in the oxylipin score plot from the blue to the red zone seem to replicate the movements of patients from COVID-19 wards to the ICUs, however this is not reflected in the cytokine score plot. Unfortunately, samples collected on different days from patients moving from the ICUs to the COVID-19 wards were not available in the biobank.

Figure 1a shows that PC1 scores provide the main contribution to the separation of the two sample classes, though PC2 also contributes. To understand which oxylipins differentiate samples, the most discriminant direction was computed as Fisher’s canonical variable (full black line), whose loadings (more precisely their absolute values) are representative of oxylipin contributions to discrimination (Figure 2). The variables with positive and negative weights in Figure 2 were over-expressed in ICU and COVID-19 samples, respectively.

**Figure 2.**
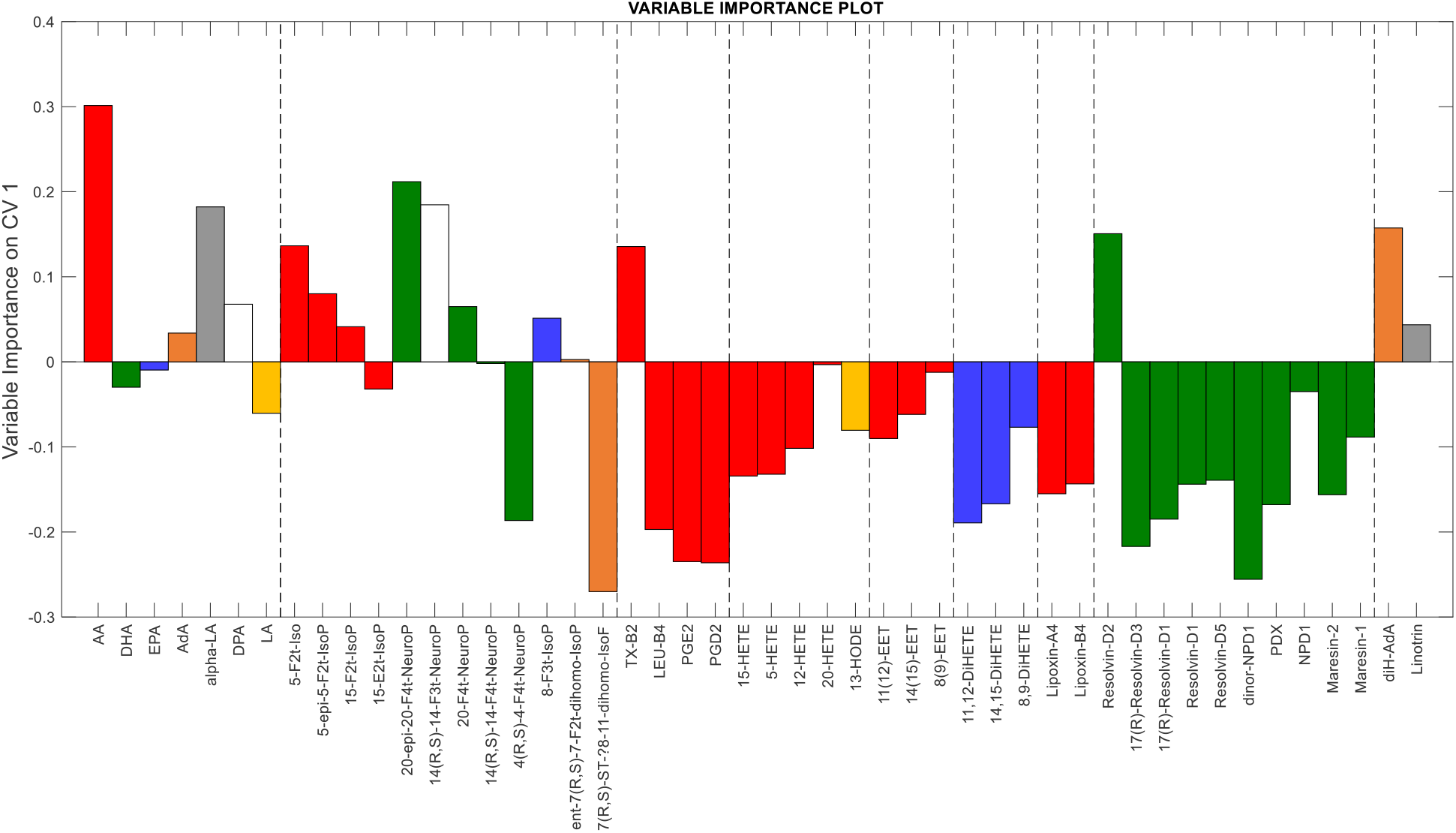
Loadings of Fisher’s canonical variable indicating the importance of the input variables in discriminating between the two classes of samples. Colours refer to different PUFAs as oxylipin precursors: red – arachidonic acid (AA); green –docosahexanoic acid (DHA); blue – eicosapentaenoic acid (EPA); orange – adrenic acid (AdA), grey – alpha-linolenic acid (alpha-LA), white – docosapentaenoic acid (DPA); yellow – linoleic acid (LA).

Samples from ICU patients were characterized by higher concentrations of most PUFAs and isoprostanoids, a group of oxylipins produced by the non-enzymatic peroxidation of membrane PUFAs, such as arachidonic acid (AA), docosahexaenoic acid (DHA), and eicosapentaenoic acid (EPA). Interestingly, thromboxane B_2_ (TX-B_2_), which is representative of the TX-A_2_ synthesis through platelet COX-1 and endothelial COX-1, and COX-2, was over-expressed in these samples. On the other hand, COVID-19 ward samples showed higher concentrations of most oxylipins of enzymatic origin: prostanoids, hydroxy-eicosatetraenoic acid (HETEs; HODE), dihydroxyeicosatetraenoic acid (DiHETEs) epoxy-eicosatrienoic acid (EETs), lipoxins, and SPMs (resolvins, protectins, maresin). Only the multivariate analysis of oxylipins provides such a clear separation between the two classes of samples, whereas individual variables exhibit a lower discriminant capability.

To further confirm the classification capacity of the full oxylipin set, a multivariate ROC curve was obtained by the UNEQ class-modelling strategy (Figure 3). The multivariate model was computed using the four lowest-order PCs as the input variables and COVID-19 samples as the target class. The resulting area under the curve (AUC, 0·92) exceeded all the AUC outcomes obtained for individual oxylipins (some univariate ROC curves are reported in **Errore. L’origine riferimento non è stata trovata**.).

**Figure 3.**
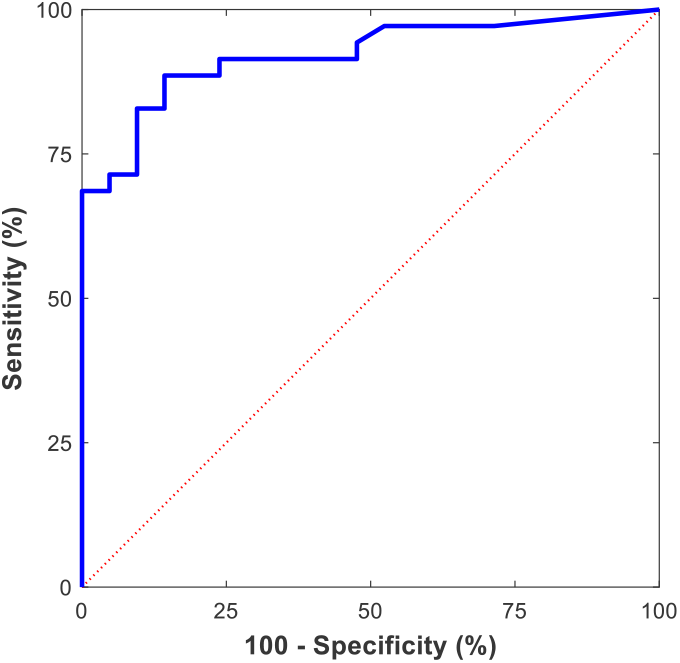
Multivariate ROC curve obtained from the UNEQ class model computed for the COVID-19 ward class with the four lowest-order PCs (AUC = 0·92).

## Discussion

Multivariate analysis of oxylipin plasma levels enabled the correct classification of COVID-19 and ICU samples, whereas separation was not achieved by processing cytokine values. The potential predictive value of this finding needs to be confirmed by prospective studies. However, the fact that many borderline samples were collected from COVID-19 ward patients that were about to be transferred to ICUs suggests that sample differences were not a trivial consequence of different treatments received from patients in different wards. In addition, the analysis of the relative contributions of individual oxylipins to the classification of the two groups fits well with our present understanding of COVID-19 and provides new insights into its pathophysiology.

ICU patients showed higher levels of pro-inflammatory isoprostanoids produced from the non-enzymatic oxidation of PUFAs such as AA, DHA and EPA, and a selective deficiency of oxylipins such as prostaglandins, leukotrienes, anti-inflammatory, and pro-resolving oxylipins originating from their enzymatic conversion. Unusually, the isoprostanoid 4(*R,S*)-4-F4t-neuroprostane was down-regulated in ICU patients. This lipid mediator prevents oxidation of the RyR ryanodine receptors, a family of Ca^2+^ release channels that controls the intracellular calcium exchange and whose oxidation may cause leaks of Ca^2+^ and lead to heart failure, pulmonary insufficiency and cognitive dysfunction in COVID-19.^29^ Furthermore, TX-B_2_ was the only prostanoid with an enzymatic origin over-expressed in ICU patients, which agrees with the need for heparin therapy in these patients to prevent microcirculatory damage.

Inflammation is a highly coordinated transcriptional process whose evolution towards resolution or persistence depends on a dynamic balance between pro- and anti-inflammatory mediators. In a controlled inflammatory process, immune tissue-resident cells activate processes producing chemokines and cytokines. Endothelial cells then respond thereby facilitating the recruitment of neutrophils (first) and monocytes (at a later stage), which differentiate into pro-inflammatory M1 macrophages (Figure 4).^30^ Pro-inflammatory cytokines activate the phospholipase A2 enzymes, which cause the release from membranes of AA, which is then converted into prostanoids or leukotrienes by cell-specific enzymatic activities. This phase is characterized by a prevalence of COX-2, both in neutrophils and M1-macrophages, and 5-LOX activity, above all in neutrophils. COX-2 and 5-LOX are responsible for the production of prostaglandins (PGD_2_ and PGE_2_) and leukotrienes (LTs), respectively.^31^

**Figure 4.**
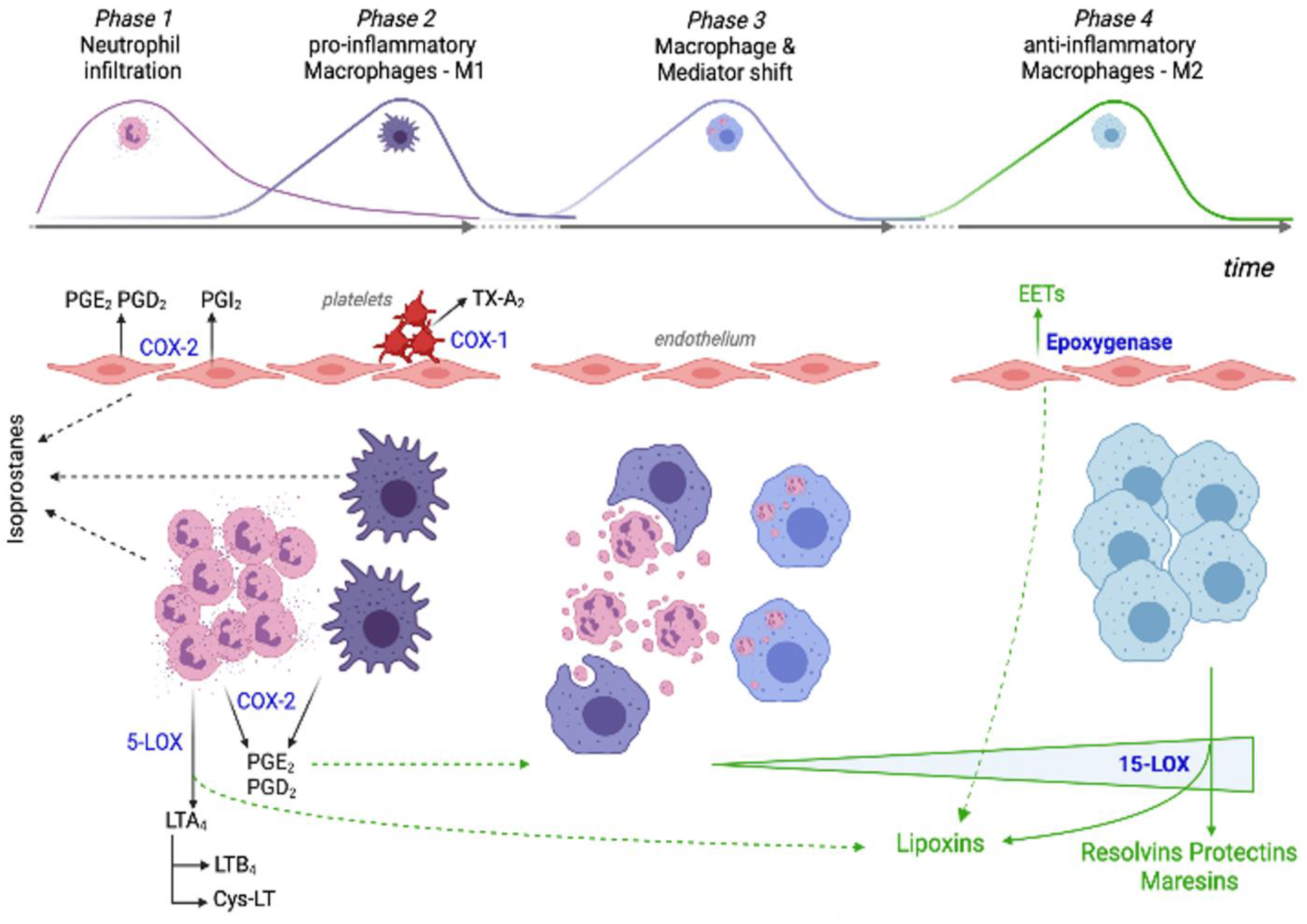
Cellular and oxylipin interplay during the evolution of an inflammatory process. Phase 1 and Phase 2: rapid neutrophil and delayed monocyte extravasation in response to cytokines produced by activated immune tissue-resident cells. Pro-inflammatory oxylipins are produced mainly by neutrophils, M1-macrophages, activated endothelial cells and platelets. Phase 3: neutrophil apoptotic bodies and prostaglandins promote the macrophage shift towards a resolution-phase function. Phase 4: inflammation resolution is promoted by increasing production of the specialized pro-resolving mediators. PGE_2_: prostaglandin E_2_; PGD_2_: prostaglandin D_2_; PGI_2_: prostacyclin I_2_; TX-A_2_: thromboxane A_2_; LTA_4_: leukotriene A_4_; LTB_4_: leukotriene B_4_; Cys-LT: cysteine-leukotrienes; EETs: epoxyeicosatrienoic acids; COX-1: cyclooxygenase-1; COX-2: cyclooxygenase-1; 5-LOX: 5-lipoxygenase; 15-LOX: 15-lipoxygenase.

The switch of lipid mediators from prostanoids to lipoxins and SPMs (resolvins, protectins, maresins) is critical for inflammation resolution.^32^ In fact, PGE_2_ facilitates the transformation of pro-inflammatory M1 into anti-inflammatory M2-macrophages characterized by the up-regulation of the 15-LOX enzyme, which is primarily involved in the synthesis of SPMs.^33^ PGE_2_ also helps to switch the pro-inflammatory (e.g. TNF-α, IL-1β and IL-6) into the anti-inflammatory interleukins synthetized by M2-macrophages (e.g. IL-10).^34^ The intermediate products of 5-LOX and 15-LOX activities (5-HPTEs and 5-HETE; 15-HPTE and 15-HETE) are also precursors for the biosynthesis of lipoxins LXA_4_ and LXB_4_, which are important agonists of the resolution of inflammation since they inhibit neutrophil recruitment, stimulate vasodilation, and promote efferocytosis.^34^

Together with prostanoids, neutrophils play a key role in the modulation of the macrophage function through the release of apoptotic bodies and microvesicles. In fact, the phagocytosis of the microvescicles by M1 macrophages is essential to trigger their functional reprogramming into M2-macrophages (Figure 4).^35^

For these reasons, we speculate that the oxylipin pattern observed in ICU patients affected by severe COVID-19 mirrors an impaired inflammation response which is part of a prolonged and unsolvable pro-inflammatory status characterized by a relative lack of oxylipins produced from the enzymatic processing of PUFAs. The impaired production of anti-inflammatory and pro-resolving oxylipins is not a consequence of the reduced availability of PUFA precursors, which appear unchanged or even increased in ICU patients. The presence of soluble isoprostanoids in both classes of patients confirms that the availability of membrane PUFAs is not a limitation for their enzymatic processing.^36^

Much remains to be understood regarding the pathogenetic mechanism of COVID-19, however defective innate and specific immune responses are likely to be critical features.^37^ Schulte-Schrepping et al.^38^ showed an increase in dysfunctional neutrophils and monocytes in severe COVID-19 patients that seems to be in agreement with our findings.

A massive endothelial dysfunction resulting from the cytokine storm and the infiltration of SARS-CoV-2 is a further characteristic of severe COVID-19 that determines the loss of vessel barrier, the promotion of leukocyte infiltration, and the activation of platelet aggregation and coagulation.^37,39^ Interestingly, in ICU patients we found higher levels of TX-B_2_, the biological inactive catabolite of TX-A_2_ (a potent activator of platelet aggregation and thus of coagulation). This is in line with the diffused microthrombosis observed in COVID-19,^37,40^ and with the lower levels of EETs, which are mainly produced by endothelial epoxygenase and are important mediators of all pro-resolving mechanisms,^41^ including the lipid mediator class switching.^42,43^

In conclusion, this study suggests that the more severe disease in ICU patients is accompanied by an inefficient enzymatic synthesis of the anti-inflammatory oxylipins resulting in an ineffective resolution mechanism of inflammation, likely worsened by endothelial damage. This hypothesis, once supported by prospective studies, might provide a basis for the identification of early biomarkers of poor disease outcome. In addition, the possible imbalance between the production of anti-inflammatory and pro-resolving oxylipins in ICU patients might explain the poor outcome of therapies based on cytokine inhibitors, and open up new perspectives for the therapy of severe COVID-19 and, in general, of lung diseases.

## Data Availability

Data is saved with the first author and can be made available if requested. but this is not stored on any URLs.

## Contributors

DB, FM, AP and FDF designed the study. DB, TL and SG developed the method for the analysis of oxylipins, LB, TD, CO and JMG synthesized 22 oxylipins needed for calibration that were not commercially available, DB, FV and AB measured oxylipin levels, MF measured cytokine levels whereas LM performed virological tests. PO performed data analysis, MF, AC and AP provided biological and clinical interpretation of data. DB, MF, PO and FDF drafted the paper, all the authors revised the paper.

## Declaration of interests

The authors declare no competing interests.

## Data sharing

All de-identified data will be shared upon approval from the IMPRE-COVID-19 steering committee and a signed data access agreement. All requests should be sent to.

## Acknowledgments

Members of the Pisa COVID group are gratefully acknowledged for collecting the clinical information, Dr. Simone Lapi and the BMS Multispecialistic Biobank of Pisa University Hospital are acknowledged for supplying the plasma samples. The study received financial support from institutional funds of the University of Pisa.

## Notes

### Competing Interest Statement

The authors have declared no competing interest.

### Author Declarations

This reasearch has been approved by Comitato Etico Area Vasta Nord Ovest. ID: IMPRE-COVID-19

