## Supporting Info for "Plasma oxylipin levels may predict Covid-19 patient outcomes. An observational and retrospective study"

### SUPPLEMENTARY INFORMATION

##### Materials

###### Detection of SARS-CoV-2 RNA

An EXTRAblood kit (ELITechGroup, catalogue number EXTB01, United Kingdom) was employed according to the manufacturer’s instructions for the manual extraction of viral nucleic acids from plasma (or serum) samples. After extraction, purified RNA samples were screened using the SARS-CoV-2 R-Gene assay (Biomerioux, Marcy-l’Etoile, France) and the 7500 Fast instrument (Applied Biosystems).

###### Quantification of oxylipins

Commercially available oxylipins, 15-F_2t_-isoprostane, 15-F_2t_-isoprostane-d4, 15-E_2t_-isoprostane, 15-E_2t_-isoprostane-d4, prostaglandin E_2_, prostaglandin E_2_-d4, prostaglandin D_2_, 15-deoxy-Δ^12,14^-Prostaglandin J_2_, thromboxane B_2_, leukotriene B_4_, lipoxin A_4_, lipoxin A_4_-d5, lipoxin B_4_, resolvin E_1_, resolvin D_1_, resolvin D_1_-d5, resolvin D_2_, resolvin D_3_, resolvin D_4_, resolvin D_5_, 17(*R*)-Resolvin-D_1_, 17(*R*)-Resolvin-D_1_-d5, 17(*R*)-Resolvin-D_3_, 17(*R*)-Resolvin-D_4_, neuroprotectin D_1_, protectin DX, maresin-1, maresin-1-d5, 7-epi-maresin-1, maresin-2, 5-hydroxyeicosatetraenoic acid (HETE), 12-HETE, 15-HETE, 20-HETE, 20-HETE-d6, 8,9-DiHETE, 11,12-DiHETE, 14,15-DiHETE, 13-hydroxy-9Z,11E-octadecadienoic acid (HODE), 8,9-epoxyeicosatrienoic acid (EET), 11,12-EET, 14,15-EET, 14,15-EET-d11, adrenic acid (AdA), eicosapentaenoic acid (EPA), alpha-linolenic acid (ALA) docosahexanoic acid (DHA), arachidonic acid (AA), docosapentaenoic acid (DPA), linoleic acid (LA) and LA-d4 (purity ≥99%) were from Cayman Chemical (Michigan, USA). Not commercially available oxylipins, i.e. 5-F_2t_-isoprostane, 5-*epi*-5-F_2t_-isoprostane, 8-F_3t_-isoprostane, 8-*epi*-8-F_3t_-isoprostane, 18-F_3t_-isoprostane, 20-F_4t_-neuroprostane, 20-*epi*-20-F_4t_-neuroprostane, 10-F_4t_-neuroprostane-d4, 10-*epi*-10-F_4t_-neuroprostane-d4, 14(*R,S*)-14-F_4t_-neuroprostane, 14(*R,S*)-14-F_3t_-neuroprostane, 4(*R,S*)-4-F_4t_-neuroprostane, C21-15-F_2t_-isoprostane, tetranor-NPD_1_, dinor-NPD_1_, 7(*R,S*)-ST-Δ^8^-11-dihomo-isofuran, ent-7(*R,S*)-7-F_2t_-dihomo-isoprostane, 17-F_2t_-dihomo-isoprostane, linotrin, diH_n-3_-DPA, diH_n-6_-DPA and diH-AdA were synthesized at the Institut des Biomolecules Max Mousseron (IBMM) (Montpellier, France), according to procedures reported elsewhere.^44–46^ All the solutions and plasma samples were stored in sterile polypropylene containers from Eppendorf (Milan, Italy). Phenex™-RC syringe filters (0·2 μm regenerate cellulose, 15 mm of diameter) were from Phenomenex (California, USA). A VELP Scientifica ZX4 Advanced Vortex Mixer (Usmate, Italy) and a Hermle Z-326 K Centrifuge, (Wehingen, Germany) were used for sample vortex-mixing and centrifugation, respectively. The removable needle micro-extraction by packed sorbent (MEPS) 250 μL syringe for HTA 300APlus (Thermo Scientific & Varian 8400 systems) and MEPS silica-C18 Barrel Insert and Needles (BINs) were purchased from SGE Analytical Science (Melbourne, Australia). The automated HT4000 Series Sample Prep workstation was purchased from HTA S.R.L. (Brescia, Italy). The UHPLC-MS/MS analysis was performed through an Agilent 1290 Infinity II LC system coupled to an Agilent 6495 triple quadrupole mass spectrometer detector. The chromatographic separation was carried out using a Polaris 3 C-18 reverse-phase column (50 × 4·6 mm, 3 μm) coupled to a guard column (5 × 4·6 mm, 3 μm), both from Agilent Technologies (Santa Clara, USA). Water, acetonitrile, and methanol were purchased from Sigma Aldrich (Milan, Italy) at LC/MS grade.

###### Quantitation of cytokines

IL-6 was quantified by Human IL-6 Instant ELISA Kit (Invitrogen, ThermoFisher Scientific), IL-1β by Human IL-1β Instant ELISA (eBioscience, Affimetryx), IL-10 by Human IL-10 Instant ELISA Kit (Invitrogen, ThermoFisher Scientific), TNF-α by Human TNF-α Quantikine® ELISA Kit (R&D System, Minneapolis, Canada), CCL2 by Human CCL2/MCP-1 Quantikine® ELISA Kit (R&D System, bio-techne), and GM-CSF by Human GM-CSF Instant ELISA kit (Invitrogen, ThermoFisher Scientific).

| **Compound** | **Internal standard** | **Quantifier transition (Q)**  **(precursor ion →**  **product ion**  **(CE, V))** | **Qualifier transition (q)**  **(precursor ion ->**  **product ion**  **(CE, V))** | **LOD**  **(pg/mL, *ng/mL)** |
| --- | --- | --- | --- | --- |
| ***PUFAs*** | | | | |
| AA | LA-d4 | 303 → 259 (12) | 303 → 285 (28) | 140^*^ |
| AdA | LA-d4 | 331 → 331 (1) |  | 60^*^ |
| DHA | LA-d4 | 327 → 283 (8) | 327 **→** 229 (10) | 100^*^ |
| DPA | LA-d4 | 329 → 285 (12) | 329 **→** 231 (8) | 50^*^ |
| EPA | LA-d4 | 301 → 257 (12) | 301 **→** 203 (16) | 150^*^ |
| LA | LA-d4 | 279 → 279 (1) |  | 200^*^ |
| alpha-LA | LA-d4 | 277 → 277 (1) |  | 90^*^ |
| ***Isoprostanoids (IsoPs, NeuroPs, dihomo-isoPs), isofurans (IsoFs) and prostanoids (prostaglandins (PGs), leukotrienes (LEUs), thromboxanes (TXs))*** | | | | |
| 5-F_2t_-IsoP | 15-F_2t_-IsoP-d4 | 353 → 309 (20) | 353 → 115 (31) | 15 |
| 5-*epi*-5-F_2t_-IsoP | 15-F_2t_-IsoP-d4 | 353 → 309 (20) | 353 → 115 (31) | 15 |
| 15-F_2t_-IsoP | 15-F_2t_-IsoP-d4 | 353 → 193 (28) | 353 → 309 (20) | 10 |
| 8-F_3t_-IsoP | C21 15-F_2t_-IsoP | 351 → 127 (28) | 351→ 155 (28) | 5 |
| 8-*epi*-8-F_3t_-IsoP^֍^ | C21 15-F_2t_-IsoP | 351 → 127 (28) | 351→ 155 (28) | 5 |
| 18-F_3t_-IsoP^֍^ | C21 15-F_2t_-IsoP | 351 → 233 (28) | 351→ 289 (28) | 10 |
| 15-E_2t_-IsoP | 15-E_2t_-IsoP-d4 | 351 → 315 (8) | 351→ 271 (28) | 10 |
| 4(*R,S*)-4-F_4t_-NeuroP | 10- F_4t_-NeuroP- d4 | 377 → 271 (25) | 377 → 101 (25) | 15 |
| 20-F_4t_-NeuroP | 10- F_4t_-NeuroP- d4 | 377 → 239 (27) | 377 → 323 (27) | 20 |
| 20-*epi*-20-F_4t_-NeuroP | 10-epi-F4_t_-NeuroP- d4 | 377 → 271 (28) | 377 → 315 (28) | 60 |
| 14(*R,S*)-14-F_4t_-NeuroP | 10- F_4t_-NeuroP- d4 | 377 → 161 (28) | 377 → 205 (28) | 20 |
| 14(*R,S*)-14-F_3t_-NeuroP | 10- F_4t_-NeuroP- d4 | 379 → 179 (30) | 379 → 207 (30) | 20 |
| ent-7(*R,S*)-7-F_2t_-dihomo-isoP | 17(R)-Resolvin-D_1_-d5 | 381 → 143 (34) | 381 → 319 (34) | 10 |
| 17-F_2t_-dihomo-isoP^֍^ | 17(R)-Resolvin-D_1_-d5 | 381 → 337 (31) | 381 → 237 (31) | 20 |
| 7(*R,S*)-ST-Δ^8^-11-dihomo-isoF | 17(R)-Resolvin-D_1_-d5 | 397 → 201 (36) | 397 → 143 (36) | 10 |
| PGE_2_ | PGE_2_-d4 | 351 → 315 (8) | 351→ 271 (28) | 10 |
| PGD_2_ | PGE_2_-d4 | 351 → 315 (12) | 351 → 271 (20) | 15 |
| 15-deoxy-Δ^12,14^-Prostaglandin J_2_^֍^ | PGE_2_-d4 | 315 → 271 (12) | 315 **→** 203 (24) | 10 |
| LEU-B_4_ | maresin 1-d5 | 335 → 195 (16) | 335 **→** 317 (16) | 90 |
| TX-B_2_ | 15-F_2t_-IsoP-d4 | 369 → 195 (12) | 369 **→**169 (20) | 50 |
| ***Hydroxy/dihydroxy-PUFAs*** | | | | |
| 5-HETE | 20-HETE-d6 | 319 → 115 (12) | 319 **→** 257 (12) | 460 |
| 12-HETE | 20-HETE-d6 | 319 → 179 (12) | 319 **→** 208 (12) | 170 |
| 15-HETE | 20-HETE-d6 | 319 → 219 (12) | 319 **→** 175 (16) | 180 |
| 20-HETE | 20-HETE-d6 | 319 → 289 (16) | 319 **→** 245 (16) | 75 |
| 13-HODE | 20-HETE-d6 | 295 → 277 (20) | 295 **→** 195 (16) | 1^*^ |
| 8,9-DiHETE | maresin 1-d5 | 335 → 185 (16) | 335 → 127 (24) | 10 |
| 11,12-DiHETE | maresin 1-d5 | 335 → 167 (16) | 335 → 207 (20) | 70 |
| 14,15-DiHETE | maresin 1-d5 | 335 → 317 (12) | 335 → 207 (20) | 60 |
| ***epoxy-PUFAs*** | | | | |
| 8(9)-EET | 14(15)-EET-d11 | 319 → 155 (12) | 319 **→** 257 (8) | 120 |
| 11(12)-EET | 14(15)-EET-d11 | 319 → 167 (12) | 319 **→** 179 (12) | 70 |
| 14(15)-EET | 14(15)-EET-d11 | 319 → 219 (8) | 319 **→** 257 (8) | 80 |
| ***Pro-resolving (lipoxins, resolvins, maresins, protectins)*** | | | | |
| Lipoxin-A_4_ | Lipoxin-A_4_-d5 | 351 → 115 (16) | 351 → 235 (14) | 5 |
| Lipoxin-B_4_ | Lipoxin-A_4_-d5 | 351 → 221 (16) | 351 → 233 (16) | 10 |
| Resolvin-D_1_ | Resolvin-D_1_-d5 | 375 → 141 (14) | 375 → 233 (14) | 10 |
| Resolvin-D_2_ | Resolvin-D_1_-d5 | 375 → 175 (18) | 375 → 141 (14) | 5 |
| Resolvin-D_3_^֍^ | Resolvin-D_1_-d5 | 375 → 147 (18) | 375 → 191 (18) | 10 |
| Resolvin-D_4_^֍^ | Resolvin-D_1_-d5 | 375 → 101 (16) | 375 → 357 (16) | 20 |
| Resolvin-D_5_ | maresin 1-d5 | 359 → 199 (16) | 359 → 279 (16) | 20 |
| 17(R)-Resolvin-D_1_ | 17(R)-Resolvin-D_1_-d5 | 375 → 141 (14) | 375 → 233 (14) | 10 |
| 17(R)-Resolvin-D_3_ | 17(R)-Resolvin-D_1_-d5 | 375 → 147 (18) | 375 → 191 (18) | 5 |
| 17(R)-Resolvin-D_4_^֍^ | Resolvin-D_1_-d5 | 375 → 101 (16) | 375 → 357 (16) | 20 |
| Resolvin-E_1_^֍^ | Resolvin-D_1_-d5 | 349 → 195 (16) | 349→ 205 (16) | 15 |
| Maresin-1 | maresin 1-d5 | 359 → 177 (16) | 359 → 250 (16) | 15 |
| 7-epi-maresin-1^֍^ | maresin 1-d5 | 359 → 250 (18) | 359 → 177 (18) | 20 |
| Maresin-2 | maresin 1-d5 | 359 → 221 (14) | 359 → 232 (16) | 15 |
| Neuroprotectin D_1_ | maresin 1-d5 | 359 → 206 (14) | 359 → 153 (14) | 20 |
| dinor-Neuroprotectin D_1_ | maresin 1-d5 | 333 → 315 (14) | 333 → 206 (21) | 100 |
| Tetranor-Neuroprotectin D_1_^֍^ | maresin 1-d5 | 305 → 243 (16) | 305 → 192 (16) | 150 |
| Protectin DX | maresin 1-d5 | 359 → 153 (14) | 359 → 206 (14) | 15 |
| diH-AdA | maresin 1-d5 | 363 → 345 (21) | 363 → 208 (21) | 30 |
| diH_n-3_-DPA^֍^ | maresin 1-d5 | 361 → 263 (16) | 361 → 206 (18) | 20 |
| diH_n-6_-DPA^֍^ | maresin 1-d5 | 361 → 153 (18) | 361 → 208 (21) | 30 |
| Linotrin | maresin 1-d5 | 309 → 291 (21) | 309 → 171 (21) | 20 |

Table S1. Full list of quantified oxylipins and relevant analytical parameters.

^֍^ These compounds showed levels below LODs in ≥ 50% of samples, thus were not included in the statistical analysis.

Table S2. Oxylipin concentration levels (ng/mL) in COVID-19 ward (W, n=43) and ICU (ICU, n=27) samples: minimum (Min), first quartile (Q_1_), median (Q_2_), third quartile (Q_3_), maximum (Max), p value (p) from Student’s t-test (difference between means) on log-transformed data.

|  |  | **Min** | **Q_1_** | **Q_2_** | **Q_3_** | **Max** | **p** |
| --- | --- | --- | --- | --- | --- | --- | --- |
| ***PUFAs*** |  |  |  |  |  |  |  |
| AA | W | 1457 | 8271 | 19456 | 27073 | 5025704 |  |
|  | ICU | 989 | 90074 | 207975 | 473039 | 1745917 | 5·90E-03 |
| AdA | W | 112 | 421 | 608 | 1286 | 3135 |  |
|  | ICU | 49 | 163 | 299 | 540 | 2333 | 1·59E-04 |
| DHA | W | 1370 | 5442 | 7892 | 15496 | 79644 |  |
|  | ICU | 611 | 1071 | 2227 | 4766 | 18899 | 1·33E-07 |
| DPA | W | 132 | 404 | 594 | 1248 | 6719 |  |
|  | ICU | 36 | 175 | 258 | 616 | 3332 | 3·32E-04 |
| EPA | W | 365 | 1109 | 2602 | 4578 | 58599 |  |
|  | ICU | 92 | 295 | 548 | 1031 | 6074 | 2·16E-06 |
| LA | W | 28932 | 93212 | 144014 | 233564 | 1144235 |  |
|  | ICU | 12213 | 33485 | 50050 | 81285 | 295489 | 6·96E-08 |
| alpha-LA | W | 534 | 2292 | 3308 | 6015 | 27592 |  |
|  | ICU | 706 | 1237 | 2086 | 6955 | 23402 | 2·34E-01 |
| **Isoprostanoids (IsoPs, NeuroPs, dihomo-isoPs), isofurans (IsoFs) and prostanoids (prostaglandins (PGs), leukotrienes (LEU), thromboxanes (TX))** | | | | | | | |
| 5-F_2t_-IsoP | W | 0·046 | 0·463 | 1·14 | 2·20 | 6·00 |  |
|  | ICU | 0·103 | 0·103 | 0·178 | 0·253 | 0·253 | 1·34E-01 |
| 5-*epi*-5-F_2t_-IsoP | W | 0·028 | 0·711 | 1·74 | 2·82 | 5·00 |  |
|  | ICU | 0·055 | 0·126 | 0·269 | 0·641 | 1·31 | 3·80E-03 |
| 15-F_2t_-IsoP | W | 0·008 | 0·035 | 0·157 | 0·295 | 0·491 |  |
|  | ICU | 0·013 | 0·028 | 0·069 | 0·219 | 1·61 | 4·14E-01 |
| 8-F_3t_-IsoP | W | 0·003 | 0·010 | 0·013 | 0·023 | 0·095 |  |
|  | ICU | 0·002 | 0·005 | 0·008 | 0·014 | 0·016 | 8·25E-03 |
| 15-E_2t_-IsoP | W | 0·006 | 0·173 | 0·334 | 0·514 | 2·03 |  |
|  | ICU | 0·002 | 0·006 | 0·014 | 0·037 | 0·201 | 9·50E-11 |
| 4(*R,S*)-4-F_4t_-NeuroP | W | 0·007 | 0·092 | 0·238 | 0·594 | 1·61 |  |
|  | ICU | 0·005 | 0·020 | 0·026 | 0·074 | 0·263 | 1·39E-05 |
| 20-F_4t_-NeuroP | W | 0·031 | 0·174 | 0·251 | 0·513 | 1·15 |  |
|  | ICU | 0·021 | 0·036 | 0·068 | 0·116 | 0·149 | 9·08E-05 |
| 20-*epi*-20-F_4t_-NeuroP | W | 0·055 | 0·240 | 0·311 | 0·548 | 1·74 |  |
|  | ICU | 0·065 | 0·195 | 0·512 | 0·795 | 2·46 | 3·76E-01 |
| 14(*R,S*)-14-F_4t_-NeuroP | W | 0·019 | 0·236 | 0·532 | 0·886 | 1·75 |  |
|  | ICU | 0·037 | 0·109 | 0·200 | 0·308 | 0·724 | 2·67E-02 |
| 14(*R,S*)-14-F_3t_-NeuroP | W | 0·004 | 0·074 | 0·144 | 0·337 | 0·695 |  |
|  | ICU | 0·007 | 0·025 | 0·038 | 0·078 | 0·191 | 1·99E-03 |
| ent-7(*R,S*)-7-F_2t_-dihomo-IsoP | W | 0·006 | 0·020 | 0·033 | 0·065 | 3·78 |  |
|  | ICU | 0·061 | 0·082 | 0·099 | 0·147 | 0·183 | 1·89E-04 |
| 7(*R,S*)-ST-Δ^8^-11-dihomo-IsoF | W | 0·133 | 1·17 | 1·65 | 2·22 | 5·33 |  |
|  | ICU | 0·061 | 0·157 | 0·313 | 1·58 | 10·8 | 2·91E-03 |
| PGE_2_ | W | 0·018 | 0·192 | 0·486 | 0·912 | 1·70 |  |
|  | ICU | 0·009 | 0·016 | 0·025 | 0·106 | 0·567 | 8·14E-10 |
| PGD_2_ | W | 0·016 | 0·120 | 0·464 | 2·24 | 13·4 |  |
|  | ICU | 0·011 | 0·021 | 0·036 | 0·083 | 0·346 | 8·61E-08 |
| LEU-B_4_ | W | 0·089 | 9·54 | 58·3 | 70·6 | 221 |  |
|  | ICU | 0·054 | 0·187 | 0·406 | 2·11 | 31·8 | 3·24E-10 |
| TX-B_2_ | W | 0·045 | 0·174 | 0·283 | 0·595 | 1·97 |  |
|  | ICU | 0·019 | 0·039 | 0·324 | 1·65 | 4·79 | 9·93E-01 |
| **Hydroxy/dihydroxy-PUFAs** | | | | | | | |
| 5-HETE | W | 0·292 | 2·32 | 49·1 | 139 | 539 |  |
|  | ICU | 0·067 | 0·419 | 0·683 | 3·36 | 45·6 | 6·34E-09 |
| 12-HETE | W | 0·593 | 3·27 | 25·4 | 62·8 | 481 |  |
|  | ICU | 0·078 | 0·918 | 1·90 | 10·6 | 20·5 | 1·37E-05 |
| 15-HETE | W | 0·363 | 2·56 | 24·7 | 82·8 | 438 |  |
|  | ICU | 0·080 | 0·562 | 0·861 | 4·51 | 21·5 | 1·37E-08 |
| 20-HETE | W | 0·049 | 0·090 | 0·136 | 0·199 | 0·891 |  |
|  | ICU | 0·019 | 0·064 | 0·103 | 0·166 | 0·379 | 1·95E-02 |
| 13-HODE | W | 19·6 | 71·3 | 229 | 517 | 1219 |  |
|  | ICU | 8·35 | 24·3 | 40·0 | 93·6 | 628 | 1·27E-05 |
| 8,9-DiHETE | W | 0·009 | 0·029 | 0·049 | 0·066 | 34·8 |  |
|  | ICU | 0·001 | 0·007 | 0·011 | 0·033 | 0·482 | 3·19E-04 |
| 11,12-DiHETE | W | 0·006 | 0·146 | 0·341 | 0·415 | 1·15 |  |
|  | ICU | 0·005 | 0·008 | 0·012 | 0·040 | 0·174 | 5·59E-10 |
| 14,15-DiHETE | W | 0·007 | 0·016 | 0·030 | 0·055 | 0·200 |  |
|  | ICU | 0·004 | 0·009 | 0·014 | 0·029 | 0·155 | 1·29E-02 |
| **Epoxy-PUFAs** | | | | | | | |
| 8(9)-EET | W | 0·073 | 0·258 | 0·712 | 1·61 | 8·18 |  |
|  | ICU | 0·064 | 0·184 | 0·235 | 0·474 | 2·45 | 1·86E-04 |
| 11(12)-EET | W | 0·046 | 0·121 | 0·264 | 0·510 | 3·18 |  |
|  | ICU | 0·029 | 0·060 | 0·095 | 0·149 | 0·291 | 5·50E-07 |
| 14(15)-EET | W | 0·054 | 0·162 | 0·517 | 1·61 | 4·43 |  |
|  | ICU | 0·068 | 0·088 | 0·169 | 0·214 | 0·778 | 4·59E-06 |
| **Pro-resolving (lipoxins, resolvins, maresins, protectins)** | | | | | | | |
| Lipoxin-A_4_ | W | 0 | 0·012 | 0·038 | 0·110 | 0·588 |  |
|  | ICU | 0·001 | 0·005 | 0·012 | 0·030 | 0·173 | 1·10E-01 |
| Lipoxin-B_4_ | W | 0·016 | 0·384 | 1·07 | 1·89 | 2·90 |  |
|  | ICU | 0·016 | 0·022 | 0·098 | 0·219 | 1·70 | 1·87E-04 |
| Resolvin-D_1_ | W | 0·002 | 0·031 | 0·170 | 0·482 | 1·78 |  |
|  | ICU | 0·002 | 0·012 | 0·022 | 0·044 | 0·641 | 5·02E-05 |
| Resolvin-D_2_ | W | 0·003 | 0·079 | 0·178 | 1·02 | 2·97 |  |
|  | ICU | 0·005 | 0·015 | 0·033 | 0·079 | 0·341 | 1·50E-02 |
| Resolvin-D_5_ | W | 0·021 | 0·632 | 1·73 | 3·95 | 31·0 |  |
|  | ICU | 0·005 | 0·038 | 0·091 | 0·489 | 3·15 | 3·88E-08 |
| 17(*R*)-Resolvin-D_1_ | W | 0·010 | 0·213 | 0·385 | 0·529 | 1·65 |  |
|  | ICU | 0·001 | 0·010 | 0·024 | 0·070 | 0·931 | 1·79E-07 |
| 17(*R*)-Resolvin-D_3_ | W | 0·003 | 0·195 | 0·286 | 0·390 | 0·911 |  |
|  | ICU | 0·001 | 0·006 | 0·024 | 0·072 | 0·242 | 5·93E-08 |
| Maresin-1 | W | 0·108 | 1·08 | 2·31 | 3·76 | 33·0 |  |
|  | ICU | 0·064 | 0·106 | 0·503 | 0·697 | 3·31 | 6·84E-04 |
| Maresin-2 | W | 0·010 | 0·137 | 0·370 | 1·18 | 19·1 |  |
|  | ICU | 0·011 | 0·017 | 0·025 | 0·073 | 0·452 | 1·31E-08 |
| Neuroprotectin D_1_ | W | 0·041 | 1·89 | 5·20 | 8·16 | 66·7 |  |
|  | ICU | 0·010 | 0·027 | 0·049 | 0·255 | 0·341 | 1·15E-08 |
| dinor-Neuroprotectin D_1_ | W | 0·096 | 2·04 | 9·65 | 20·4 | 116 |  |
|  | ICU | 0·058 | 0·146 | 0·275 | 0·503 | 2·90 | 6·60E-12 |
| Protectin DX | W | 0·010 | 0·976 | 3·21 | 5·96 | 41·9 |  |
|  | ICU | 0·009 | 0·037 | 0·086 | 0·707 | 5·26 | 2·80E-08 |
| diH-AdA | W | 0·008 | 0·143 | 0·517 | 1·11 | 16·2 |  |
|  | ICU | 0·008 | 0·044 | 0·066 | 0·084 | 0·088 | 7·07E-03 |
| Linotrin | W | 0·016 | 0·042 | 0·102 | 0·173 | 0·580 |  |
|  | ICU | 0·007 | 0·021 | 0·049 | 0·115 | 0·497 | 2·23E-02 |

Table S3. Cytokine concentration levels (pg/mL) in COVID-19 ward (W, n=43) and ICU (ICU, n=27) samples: minimum (Min), first quartile (Q1), median (Q2), third quartile (Q3), maximum (Max), p value (p) from Student’s t-test (difference between means) on log-transformed data.

|  |  | **Min** | **Q_1_** | **Q_2_** | **Q_3_** | **Max** | **p** |
| --- | --- | --- | --- | --- | --- | --- | --- |
| IL-6 | W | 2·7 | 12·7 | 20·1 | 42·7 | 602 |  |
|  | ICU | 1·3 | 15·8 | 53·0 | 129 | 614 | 0·332 |
| IL-1β | W | 0·2 | 0·7 | 0·9 | 1·5 | 31·0 |  |
|  | ICU | 0·2 | 0·7 | 1·2 | 2·1 | 16·4 | 0·363 |
| IL-10 | W | 0·2 | 1·0 | 2·7 | 3·4 | 324 |  |
|  | ICU | 0·4 | 1·0 | 3·4 | 5·4 | 15·8 | 0·949 |
| TNF-α | W | 3·3 | 5·5 | 7·8 | 15·2 | 32·5 |  |
|  | ICU | 0·6 | 2·8 | 8·5 | 20·0 | 50·3 | 0·363 |
| CCL2 | W | 175 | 357 | 600 | 910 | 3950 |  |
|  | ICU | 172 | 411 | 630 | 1852 | 3971 | 0·254 |

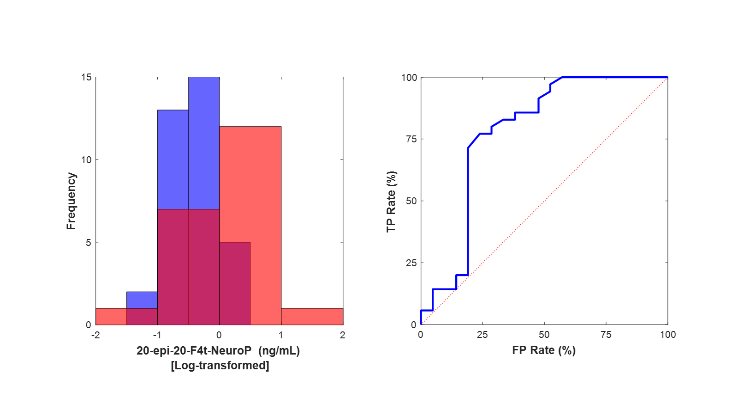

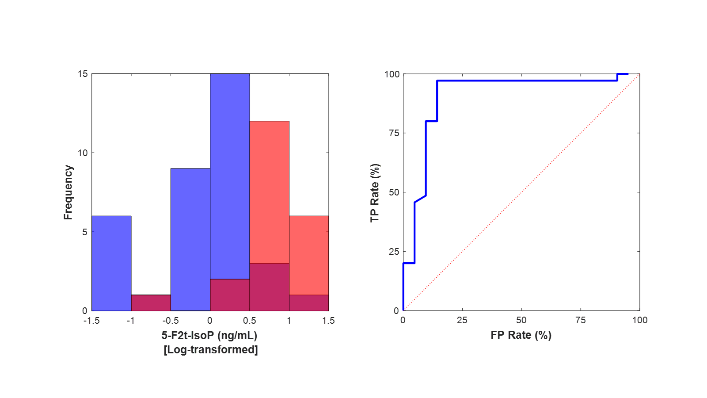

Figure S1. ROC curves of the best performing oxylipins in the classification of samples: 20-epi-20-F_4t_-NeuroP (top, AUC= 0·78) and 5-F_2t_-IsoP (bottom, AUC = 0·88). FP = false positives; TP = true positives.
